# Mismatch: A comparative study of vitamin D status in British-Bangladeshi migrants

**DOI:** 10.1101/2020.12.04.20244111

**Authors:** Nicholas Smith, Lynnette Leidy Sievert, Shanthi Muttukrishna, Khurshida Begum, Lorna Murphy, Taniya Sharmeen, Richard Gunu, Osul Chowdhury, Gillian R Bentley

## Abstract

**Background and objectives:** Low levels of vitamin D among dark-skinned migrants to northern latitudes and increased risks for associated pathologies illustrate an evolutionary mismatch between an environment of high ultraviolet (UV) radiation to which such migrants are adapted and the low-UV environment to which they migrate. Recently, low levels of vitamin D have also been associated with higher risks for contracting COVID-19. South Asians in the UK have higher risk for low vitamin D levels. In this study, we assessed vitamin D status of British-Bangladeshi migrants compared to white British residents and Bangladeshis still living in Bangladesh (‘sedentees’).

**Methodology:** The cross-sectional study compared vitamin D levels among 149 women aged 35-59, comprising British-Bangladeshi migrants (n=50), white UK neighbors (n=54) and Bangladeshi sedentees (n=45). Analyses comprised multivariate models to assess serum levels of 25-hydroxyvitamin D (25(OH)D), and associations with anthropometric, lifestyle, health and migration factors.

**Results:** Vitamin D levels in Bangladeshi migrants were very low: mean 25(OH)D = 32.2nmol/L ± 13.0, with 29% of migrants classified as deficient (<25nmol/L) and 94% deficient or insufficient (≤50nmol/L). Mean levels of vitamin D were significantly lower among British-Bangladeshis compared to Bangladeshi sedentees (50.9nmol/L ± 13.3), presumably due to less exposure to sunlight following migration; levels were also lower than in white British women (55.3nmol/L ± 20.9). Lower levels of vitamin D were associated with increased body mass index and low iron status.

**Conclusions and implications:** Recommending supplements to Bangladeshi migrants could prevent potentially adverse health outcomes associated with vitamin D deficiency.

**Lay summary:** Vitamin D deficiency is one example of mismatch between an evolved trait and novel environments. Here we compare vitamin D status of dark-skinned British-Bangladeshi migrants in the UK to Bangladeshis in Bangladesh and white British individuals. Migrants had lower levels of vitamin D and are at risk for associated pathologies.

## Introduction

Human skin color evolved over several thousand years as our ancestors migrated from warmer, lower latitudes to northern climes where there was significantly less sunlight [1]. Depigmented skin is thought to have evolved over thousands of years, primarily to facilitate vitamin D absorption in the new geographic regions to where humans slowly migrated. The phenomenon of more rapid migration only occurred when humans evolved the technological capacity to travel quickly over vast distances. In evolutionary medicine, the concept of “mismatch” refers to pathologies that occur when traits that evolved under one set of conditions no longer fit the novel environments in which humans find themselves [2,3]. Vitamin D deficiency falls into this category of evolutionary explanation because, following rapid migration, darkly pigmented skin can be detrimental in northern latitudes leading to vitamin D deficiency with a variety of health consequences [4,5]. This deficiency can be exacerbated where cultural practices, such as clothing and cloistering, also prevent exposure to sunlight [4,6]. Conversely, the migration of depigmented individuals to southern latitudes has led to a steep rise in skin cancer, especially where individuals fail to protect themselves from ultraviolet (UV) exposure [2].

Up to a billion people worldwide are currently estimated to have low vitamin D levels, making it one of the most common micronutrient deficiencies in humans [7,8]. Health impacts can be severe, including risk of rickets and osteomalacia [9,10]; additional health impacts are disputed in relation to cancers [11,12], dementia [13] and cardiovascular diseases [12,14]. Effects are most pronounced in cases of clinical vitamin D ‘deficiency’ (serum concentrations of 25-hydroxyvitamin D (25(OH)D) <25nmol/L), but even ‘insufficiency’ (concentrations ≤50nmol/L) can produce clinical features such as lower bone density [15], non-specific muscle and joint pain [16,17], increased risk of falls and fractures [18] and weaker muscle strength [16,19]. Some of these have been shown to be reversible with vitamin D supplementation, such as bone density, risk of falls and fracture risk [18,20].

More recently, low vitamin D has been linked to risks for greater morbidity and mortality for COVID-19, although this association remains controversial [21]. In a recent study of health workers in Israel, levels of plasma 25(OH)D were significantly lower among those who were infected with COVID-19 [22]. However other studies have found no association between 25(OH)D levels and risks for COVID-19 after controlling for socioeconomic, demographic and other predisposing health risk factors [23]. Previous meta-analyses have shown that vitamin D supplementation can help to prevent respiratory diseases, especially in deficient individuals [24]; it could be that this is also true for COVID-19.

In the UK, vitamin D deficiency is a well-recognized public health issue, present in just over a quarter of adults from January to March, and in only 4% of adults from July to September [25]. Mean 25(OH)D in adults aged 19-64 varies seasonally from 36.2nmol/L (January to March) to 57.7nmol/L (July to September). A comprehensive model developed by O’Neill [26] suggests that vitamin D is lowest in white British between November and April, and highest from May to October, with a peak in August and a nadir in January. Other general factors associated with poor vitamin D status include being overweight [27], smoking [28], chewing betel quid (a common practice among South Asians) [29], and low iron levels [30], although with the exception of obesity, the direction of causality of these factors is not firmly established [27].

Low vitamin D is especially common in darker-skinned migrants to the UK [4,5], and is mirrored in other European countries including among Bangladeshi migrants in Greece [31], and Sri Lankans in Norway [32]. Even in their countries of origin, some South Asians are reported to be deficient in vitamin D depending on lifestyle factors presumably affecting outdoor exposures [6]. In South Asians, the associations between low 25(OH)D, rickets, and non-specific musculoskeletal problems is also well established [17,33]. Islam et al. [20] showed that supplementation with vitamin D had a positive effect on bone mineral density (BMD) in Bangladeshi women with low levels of 25(OH)D, indicating that some of the negative impacts of vitamin D deficiency can be readily reversed in this population.

The UK Department of Health guidelines currently recommend regular vitamin D supplementation for anyone with clinically defined deficiency [34]. For those with 25(OH)D levels of 25-50nmol/L, supplementation is recommended if a person is at higher risk than average for adverse consequences of vitamin D insufficiency, such as those with bone disease, the elderly, the obese and those with darker skin. Currently it is estimated that only 23-39% of British South Asians take vitamin D supplements [35].

The aim of this study was to compare vitamin D levels among: i) British-Bangladeshi migrant women; ii) white British women living in the UK; and iii) Bangladeshi women in their country of origin (‘sedentees’). We aimed to assess associations between vitamin D status and potential determinants, such as time of year, anthropometric factors, other nutrient deficiencies (such as iron), educational status, time since migration and lifestyle factors (such as tobacco use). Given the recent history of migration of Bangladeshis to the UK and the mismatch between their skin pigmentation and low UV exposure in their new environment, we hypothesized that British-Bangladeshi migrants would, on average, be deficient in vitamin D. We also hypothesized that the white British and Bangladeshi sedentee women would, on average, be sufficient in vitamin D.

## Methodology

### Participants and Recruitment

Between March, 2007, and May, 2010, 149 women aged 35-59 who were willing to undergo phlebotomy were recruited into a wider study of reproductive ageing [36,37]. They comprised: 1) 50 first-generation, Bangladeshi migrants living in London, UK and originating from Sylhet, northeast Bangladesh, 2) 54 white British women of European origin also living in London, and 3) 45 middle-class, sedentee Bangladeshis in Sylhet, Bangladesh, matched to the UK migrants for socioeconomic status and likelihood of migration. UK-based participants were recruited from community centers, advertisements in local newspapers and posters in public places (such as doctors’ surgeries and libraries). The women in Sylhet were recruited using personal networks and snowball techniques (where participant contacts help to expand recruitment networks).

Participants were screened (by email, telephone or face-to-face interviews) according to the requirements of the original study. Exclusion criteria included being pregnant or lactating, having thyroid disorders, a hysterectomy/oophorectomy, or use of exogenous hormones (such as hormone replacement therapy) in the prior three months. It was not anticipated that any of these exclusion criteria would have affected vitamin D status except for pregnancy/lactation. The white British women all had parents born either in the UK or the Republic of Ireland.

### Questionnaires

A three-part questionnaire was administered to all participants during face-to-face interviews. These were conducted in English for the white British group while, for Bangladeshi migrants and sedentees, either Bangla (the national language of Bangladesh) or Sylheti (the local dialect) was used by a researcher fluent in the relevant language (KB or TS).

The questionnaire contained information on demographics (age, religion, self-reported ethnicity, years of education), migration history (for Bangladeshi migrants, age at migration and therefore number of years in the UK), and lifestyle information (calcium and non-specific vitamin supplementation, smoking, betel nut consumption). Almost half of the sedentees (49%, n=22) and 1 migrant (2%) were born before official recording of births in Bangladesh and were, therefore, uncertain of their exact age. These participants were asked to estimate their date of birth using a 21- item event calendar (e.g., the date of the India-Pakistan war or specific natural disasters such as cyclones).

### Anthropometry

Participants were measured for their height, weight, waist circumference, hip circumference, triceps skinfold thickness and mid-upper arm circumference using standardized techniques [38]. From these measurements both the BMI and waist-to-hip ratio (WHR) were calculated.

### Serum samples

One blood sample of 5 ml was taken from each woman using methods described elsewhere [39]. All samples for the sedentee Bangladeshis were taken in March and April (because of relatively constant levels of sunshine across the entire year), while samples from the other groups were taken throughout the year. To separate the serum, samples were centrifuged, then stored at −20° C in laboratories at University College London (UCL) or the M.A.G. Osmani Medical College, Sylhet, Bangladesh. Samples from Sylhet were transported on dry ice to UCL where analyses of all samples were performed together. Both 25(OH)D and ferritin were analyzed using an electrochemiluminescence immunoassay (ECLIA) kit (Roche Molecular, Biochemicals, Mannheim, Germany). Although this method has since been superseded by liquid chromatography tandem mass spectrometry (LC/MS/MS) as the gold standard to measure 25(OH)D, it has been shown to have acceptable performance [40]. Total 25(OH)D was measured (sum of vitamin D_2_ and D_3_), hereafter referred to simply as ‘25(OH)D’. For 25(OH)D, mean intra- and inter-assay CVs were <10%, and the measuring range of the assay was 10nmol/L to 250nmol/L. For ferritin, the mean CV was <11.6%, with a measuring range from 0.5 to 2000ng/ml. Following standard procedures [41] vitamin D status was defined as follows: levels >50nmol/L are ‘sufficient’, 25-50nmol/L are ‘insufficient’, and levels <25nmol/L are ‘deficient’.

### Data Analyses

Since vitamin D status varies dramatically over the course of a year among people living in the UK due to varying levels of sunlight [25], the white British and British-Bangladeshi samples were grouped into two seasons, following O’Neill et al. [26]: from May to October when vitamin D levels tend to be highest (extended ‘summer’), and November to April when levels are normally lowest (extended ‘winter’). The majority of both groups participated in the summer (67% migrants and 70% white British). The same proportion of women were sampled in the summer and winter in each group (p=0.832, Table S2-Supplemental). Differences in serum 25(OH)D between grouped summer and winter samples were analyzed using independent t-tests.

For analyses including BMI, results were grouped into WHO categories [42] (<18.5=underweight, ≥18.5 and <25=healthy, 25-30=overweight and >30=obese), but since there were only three underweight women, BMI categories were combined into two groups: <25 (normal weight) and ≥25 (overweight and obese).

Since Bangladeshis had fewer years of education, on average, than the white British women, we defined ‘low education’ as <10 years in whites and <6 years in Bangladeshis, and ‘high education’ as ≥10 and ≥6 years in white women and Bangladeshi women, respectively. Independent t-tests were then performed to assess for differences in serum 25(OH)D between low and high education groups.

#### Univariate statistics

Chi-square analyses were performed to detect significant differences in vitamin D status (deficient, insufficient, sufficient) between each group. To assess differences between groups (white British, Bangladeshi migrant and Bangladeshi sedentee), variables that were normally distributed (age, height, weight, BMI, WHR, triceps skin fold thickness, arm circumference, 25(OH)D) were analyzed using one-way ANOVAs with Bonferroni post-hoc. Variables that were not normally distributed (ferritin, years of education) were analyzed using Kruskal-Wallis tests. For smoking and betel nut use with tobacco, results were combined into one binary variable (‘any tobacco usage’). Calcium supplements and vitamin/mineral supplements were also combined into an ‘any supplements’ variable. Chi-square tests were performed to assess differences in these variables across the three groups.

Bivariate analyses of associations between levels of vitamin D and linear variables (BMI, age, WHR, triceps skin fold thickness, arm circumference, time since migration) were performed using Pearson correlations. Independent t-tests were then performed, comparing vitamin D deficient versus not deficient, and again comparing insufficient versus sufficient. Independent t-tests were used to assess associations between BMI categories and vitamin D levels. As a covariate, ferritin status was grouped into ‘low’ and ‘adequate’ according to the recommended level set by the manufacturer of the assay (<13ng/ml for low, ≥13ng/ml for adequate). Participants with abnormally high levels of ferritin (>150ng/ml) were excluded from the analyses. Fisher’s exact tests were performed to analyze associations between ferritin status and vitamin D sufficiency status (≤50nmol/L vs >50nmol/L). Independent t-tests were performed to assess differences in mean 25(OH)D between those who used tobacco versus non-users, and those who took supplements versus no supplements.

#### Multivariate analyses

Multivariate analyses were performed using all variables that approached significance in bivariate tests (yielding a p-value <0.2). Since triceps skin fold thickness was significantly associated with vitamin D levels, but correlated with BMI, the latter was included in the analyses. To account for the fact that there was minimal seasonal variation in sunlight exposure in Bangladesh, the participants were split into two groups: Group ‘A’, comprising the white British women and Bangladeshi migrants, and Group ‘B’, the Bangladeshi sedentees. For Group A, variables included in each model were: 1) season of sampling, 2) group (i.e., white vs migrant), 3) ferritin status, 4) BMI and 5) age. For Group B, variables were: 1) ferritin status, 2) BMI and 3) age.

For both Groups A and B, three models were then analyzed. Model 1 assessed associations between the covariates and vitamin D as a continuous variable using linear regression. Models 2 and 3 used binary logistic regression: Model 2 assessed whether there was an association between the covariates mentioned above and being either deficient (i.e., <25nmol/L 25(OH)D) or not deficient. Model 3 did the same but used the higher cut-off of sufficiency (i.e., ≤50nmol/L 25(OH)D). Therefore, there are six models in total: three for group A, labelled A1, A2 and A3, and the same for group B (B1, B2, B3). Model B2 was not considered valid as there were only 2 participants in the deficient group, limiting statistical analysis.

Statistical analyses were performed using SPSS 24.0. Significance for all tests was set at p≤0.05.

### Ethical approval

Ethical approval for the study was obtained from the Institutional Review Board of UMass Amherst, and the Ethics Committees at University College London, Durham University, and the M.A.G. Osmani Medical College, Sylhet, Bangladesh.

## Results

### Sample characteristics

One participant in the Bangladeshi migrant group had a 25(OH)D level >4 times the upper assay limit, so was excluded from analyses. Therefore, the final sample included 148 participants (49 Bangladeshi migrants, 54 white British women and 45 Bangladeshi sedentees).

Sample characteristics are shown in Table 1. The mean age of all women was 46.4 years with no significant differences across the three groups. Mean BMI was higher in the migrant Bangladeshi group than the sedentees (27.1 and 24.9, respectively, p=0.049). White British women had more years of education than the other two groups (13.1 compared to 8.9 in migrants and 9.4 in sedentees, p<0.001). Mean ferritin was highest in the white women (56.3ng/ml) and lowest in the migrants (35.2ng/ml), and differed significantly between these two groups (p=0.044), but not between the sedentees and either of the other groups.

**Table 1:** Descriptive statistics of study participants.

| Table values indicate mean (SD) |  |  |  |  |  |  |  |  |
| --- | --- | --- | --- | --- | --- | --- | --- | --- |
| Physical and biochemical characteristics | White British<br>n=54 | Bangladeshi<br>migrants<br>n=49 | Bangladeshi<br>sedentees<br>n=45 | Total cohort<br>n=148 | p ANOVA<br>between<br>groups | p W-BM<sup>a</sup> | p BM-BS<sup>b</sup> | p W-BS<sup>c</sup> |
| Age (years) | 47.1 (7.0) | 45.6 (6.6) | 46.6 (6.9) | 46.4 (6.8) | 0.541 | 0.831 | 1.000 | 1.000 |
| Height (cm) | 162.4 (5.9) | 152.8 (7.0) | 150.6 (5.6) | 155.7 (8.1) | <0.001 | <0.001 | 0.289 | <0.001 |
| Weight (kg) | 67.4 (14.4) | 63.5 (10.2) | 56.6 (11.6) | 62.8 (13.0) | <0.001 | 0.319 | 0.023 | <0.001 |
| BMI (kg/m<sup>2</sup>) | 25.5 (5.0) | 27.1 (3.5) | 24.9 (4.6) | 25.8 (4.5) | 0.042 | 0.190 | 0.049 | 1.000 |
| Waist-hip ratio<sup>d</sup> | 0.78 (0.09) | 0.85 (0.06) | 0.83 (0.06) | 0.82 (0.08) | <0.001 | <0.001 | 0.779 | 0.002 |
| Triceps skin fold thickness<sup>e</sup><br>(cm) | 20.4 (6.5) | 28.8 (7.3) | 23.8 (8.8) | 24.2 (8.3) | <0.001 | <0.001 | 0.005 | 0.090 |
| Arm circumference<sup>f</sup> (cm) | 28.3 (3.7) | 30.4 (4.0) | 27.5 (4.7) | 28.8 (4.3) | 0.003 | 0.046 | 0.003 | 1.000 |
| Ferritin (ng/ml) | 56.31 (53.76) | 35.21 (36.99) | 45.76 (34.53) | 46.11 (43.86) | 0.011 | 0.044 | 0.719 | 0.686 |
| Years of education<sup>g</sup> | 13.1 (2.4) | 8.9 (3.3) | 9.4 (3.8) | 10.7 (3.7) | <0.001 | <0.001 | 1.000 | <0.001 |
| Vitamin D status: number of participants (percentage of group) |  |  |  |  |  |  |  |  |
| Deficient (<25nmol/L) | 3 (5.6%) | 14 (28.6%) | 2 (4.4%) | 19 (12.8%) | p<0.001 |  |  |  |
| Insufficient (25-50nmol/L) | 21 (38.9%) | 32 (65.3%) | 17 (37.8%) | 70 (47.3%) |  |  |  |  |
| Sufficient (>50nmol/L) | 30 (55.6%) | 3 (6.1%) | 26 (57.8%) | 59 (39.9%) |  |  |  |  |

25(OH)D, 25-hydroxyvitamin D<sub>3</sub>. p-values in **bold** indicate significant difference between the groups (p<0.05). <sup>a</sup> p-value between White British and Bangladeshi migrant groups. <sup>b</sup> p-value between Bangladeshi migrant and Bangladeshi sedentee groups <sup>c</sup> p-value between White British and Bangladeshi sedentee groups. <sup>d</sup> missing data for n=3 whites, n=1 migrant. <sup>e</sup> missing data for n=2 whites, n=1 migrant. <sup>f</sup> missing data for n=5 whites, n=1 migrant. <sup>g</sup> missing data for n=8 migrants, n=1 sedentee.

White British women were more likely to smoke (13.7% vs. no smokers among either migrants or sedentees), but many migrants and sedentees chewed betel nut quid that contained tobacco (30.6% and 48.8% respectively). There was no significant difference in calcium or vitamin supplement usage between the groups (with 6-16% of each group taking calcium supplements and 20-37% taking any other supplements). Migrant Bangladeshis were all Muslim while 36.4% of the sedentees were Hindu. These data are displayed in Table S1 – Supplemental.

### Vitamin D status

Vitamin D levels were significantly lower in migrant Bangladeshis than the other two groups: mean 25(OH)D in migrants was 32.2nmol/L ± 13.0 compared to 55.3nmol/L ± 20.9 in white British women and 50.9nmol/L ± 13.3 in sedentees (p<0.001, Figure 1). Bangladeshi migrants were therefore, on average, insufficient in vitamin D, while a significant proportion was deficient (Table 1). Most sedentees (58%) were sufficient, with only 2 individuals (4%) being deficient, while 56% of the white population in London was sufficient with likewise a very low proportion (6%) being deficient. However, among both the sedentees and white British, a large proportion was insufficient (38% and 39%, respectively).

**Figure 1:**
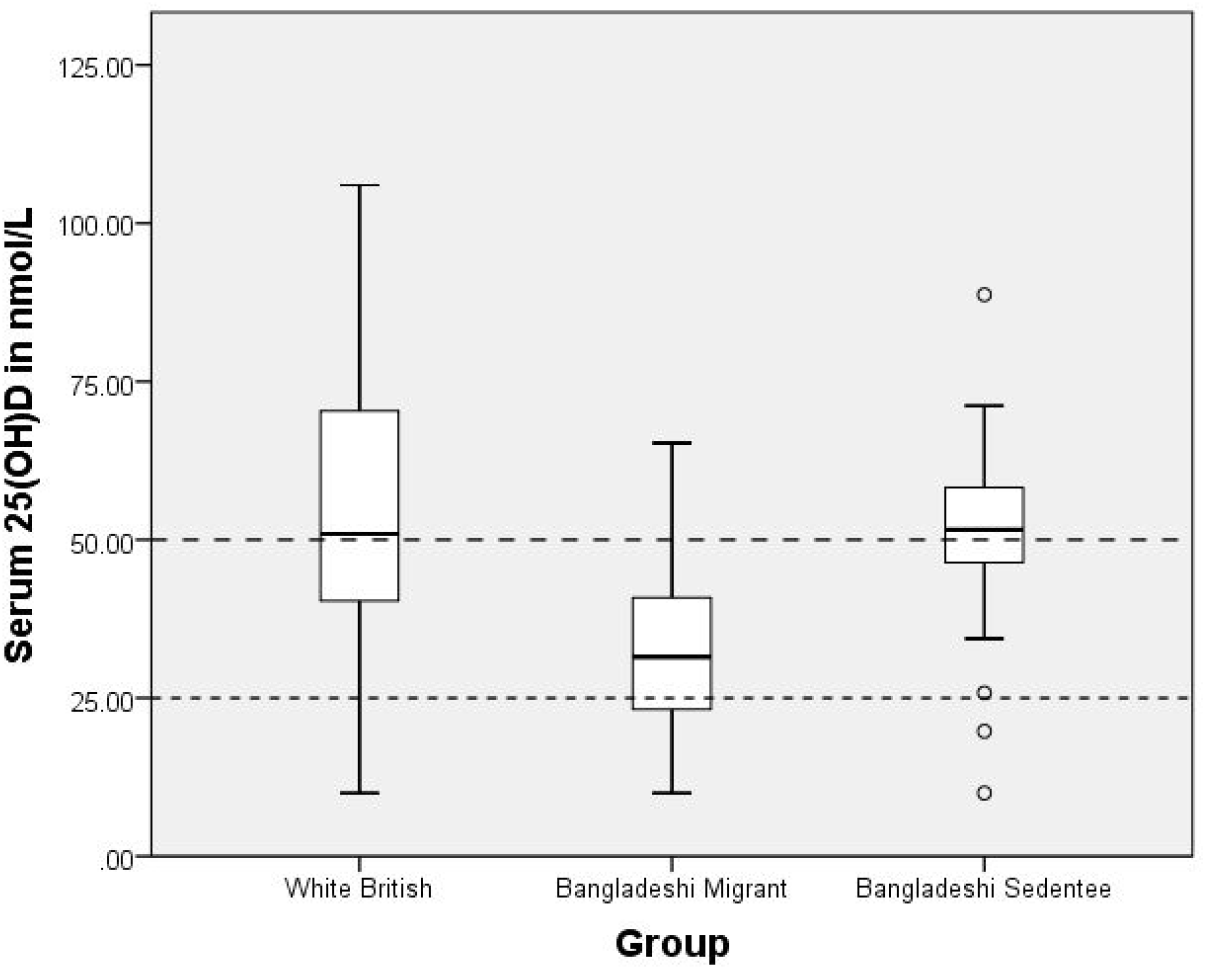
Serum 25(OH)D in each group (white British, Bangladeshi migrant and Bangladeshi sedentee). Box-plot indicates upper/lower limits, upper/lower quartiles and median, with circles for significant outliers. Lines at 25nmol/L and 50nmol/L indicate ‘deficiency’ and ‘insufficiency’, respectively. Significant difference between white British and migrants (<0.001) and between migrants and sedentees (p<0.001) but not between white British and sedentees (p=0.558).

There was a significant difference between winter and summer values in the white British group (summer mean 25(OH)D 59.9nmol/L ± 18.2; winter mean 25(OH)D 44.2nmol/L ± 23.1, p=0.010), but not in the Bangladeshi migrant group although the winter values were lower (summer mean 25(OH)D 34.6nmol/L ± 12.1; winter mean 25(OH)D 27.2nmol/L ± 13.9, p=0.061, Figure 2).

**Figure 2:**
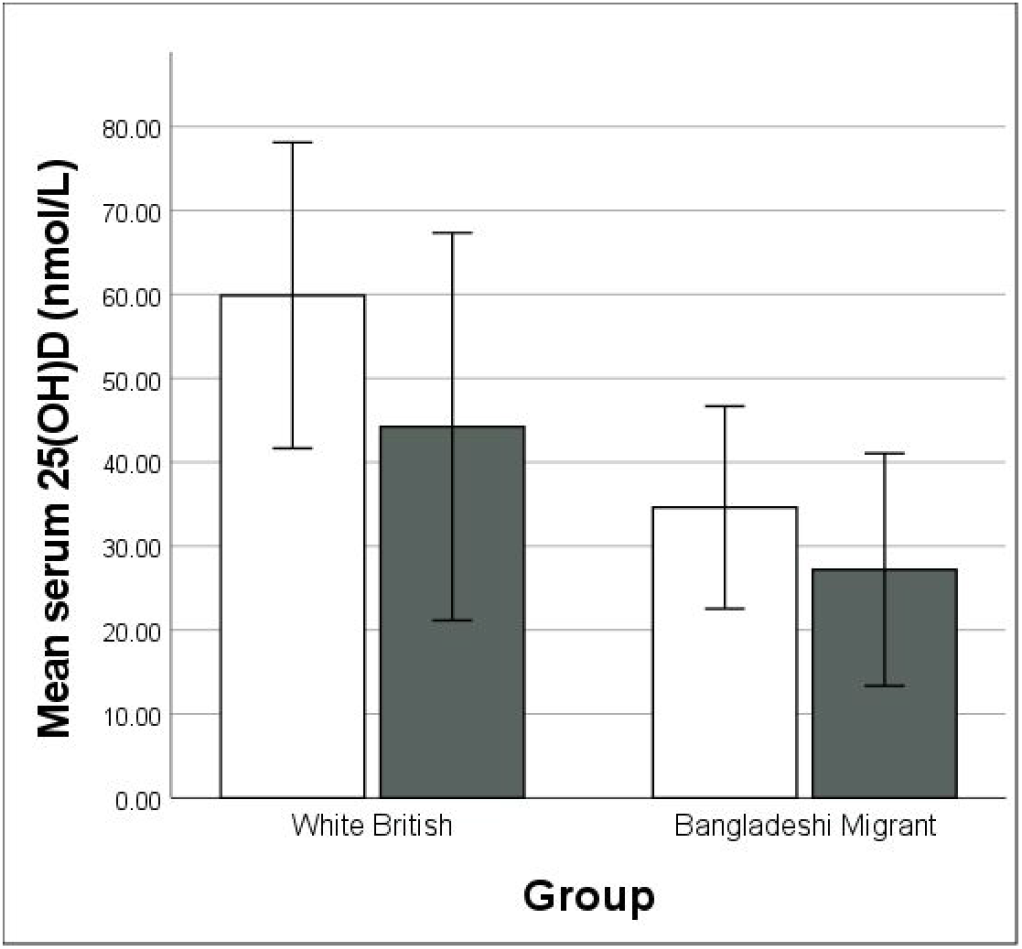
Mean serum 25(OH)D in each group in extended ‘summer’ (May-Oct) vs extended ‘winter’ (Nov-Apr). White bars (□) indicate summer, shaded bars (▪) indicate winter. Error bars indicate ± 1 SD. Note: Bangladeshi sedentee data not included in comparison as in Bangladesh, UV radiation varies less across the year, and is highest/lowest in different months of the year to the UK.

Vitamin D insufficiency was associated with low iron status (p=0.02; Table S3 - Supplemental). Vitamin D levels were also lower in overweight/obese women (BMI ≥25, mean vitamin D 43.4nmol/L) than in those with a BMI <25 (mean vitamin D 49.8nmol/L, p=0.043). Other anthropometric variables which could be expected to vary with BMI (WHR, triceps skin fold thickness) showed similar correlations with vitamin D (see Supplemental Materials, Tables S4 and S5).

There was no association between vitamin D status and age, educational status, tobacco use, non- specific supplement use, or length of time since migration (see Supplemental Materials, Table S6).

In each of the three ‘Group A’ (white and migrant) regression models that included group, age, season of sampling, BMI, and ferritin status as covariates, only group and season of sampling were significantly associated with levels of vitamin D (Table 2). In the ‘Group B’ (sedentee) models, we found no significant explanatory variables for vitamin D status (Table 3).

**Table 2:** Explanatory models for Group A (white British and Bangladeshi migrant women)

| **Model A1:**<br>**Vitamin D as a continuous variable in white British and Bangladeshi migrant women,**<br>**n=103** |  |  |  |  |
| --- | --- | --- | --- | --- |
| **Variable** | **B** | **95% CI (lower)** | **95% CI (upper)** | **p** |
| Winter season<sup>a</sup> | -11.776 | -18.997 | -4.554 | **0.002** |
| Bangladeshi migrant<sup>b</sup> | -20.696 | -27.643 | -13.748 | **<0.001** |
| Low ferritin status<sup>c</sup> | -4.579 | -13.325 | 4.167 | 0.301 |
| BMI | -0.459 | -1.222 | 0.304 | 0.236 |
| Age | 0.186 | -0.313 | 0.685 | 0.462 |
| **Model A2:**<br>**Odds ratios in whites and migrants of being deficient, n=17, vs not deficient, n=86** |  |  |  |  |
| **Variable** | **OR** | **95% CI (lower)** | **95% CI (upper)** | **p** |
| Winter season<sup>a</sup> | 0.116 | 0.031 | 0.432 | **0.001** |
| Bangladeshi migrant<sup>b</sup> | 0.141 | 0.034 | 0.586 | **0.007** |
| Low ferritin status<sup>c</sup> | 0.479 | 0.116 | 1.971 | 0.308 |
| BMI | 0.941 | 0.807 | 1.097 | 0.437 |
| Age | 0.985 | 0.901 | 1.078 | 0.746 |
| **Model A3:**<br>**Odds ratios in white British and migrants of being insufficient, n=70, vs sufficient, n=33** |  |  |  |  |
| **Variable** | **OR** | **95% CI (lower)** | **95% CI (upper)** | **p** |
| Winter season<sup>a</sup> | 0.239 | 0.068 | 0.834 | **0.025** |
| Bangladeshi migrant<sup>b</sup> | 0.061 | 0.016 | 0.234 | **<0.001** |
| Low ferritin status<sup>c</sup> | 0.412 | 0.070 | 2.413 | 0.326 |
| BMI | 0.937 | 0.837 | 1.049 | 0.260 |
| Age | 1.036 | 0.956 | 1.123 | 0.383 |

P values which reached significance (p=0.05) are shown in **bold**.
Adjusted R<sup>2</sup> for model A1: 0.352.
<sup>a</sup> reference: summer season
<sup>b</sup> reference: White British
<sup>c</sup> defined as ferritin <13ng/ml. Reference group: ferritin ≥13ng/ml

**Table 3:** Explanatory models for Group B (Bangladeshi sedentee women)

| Model B1:<br>Vitamin D as a continuous variable in Bangladeshi sedentees, n=45 |  |  |  |  |
| --- | --- | --- | --- | --- |
| Variable | B | 95% CI (lower) | 95% CI (upper) | p |
| Low ferritin status<sup>a</sup> | 0.841 | -10.680 | 12.362 | 0.884 |
| BMI | -0.319 | -1.272 | 0.635 | 0.504 |
| Age | -0.271 | -0.921 | 0.380 | 0.405 |
| Model B3:<br>Odds ratios in sedentees of being insufficient, n=19, vs sufficient, n=26 |  |  |  |  |
| Variable | OR | 95% CI (lower) | 95% CI (upper) | p |
| Low ferritin status<sup>a</sup> | 1.108 | 0.205 | 5.984 | 0.905 |
| BMI | 0.918 | 0.793 | 1.062 | 0.250 |
| Age | 1.044 | 0.947 | 1.150 | 0.387 |

Model B2 has not been included due to small sample size in the deficient group (n=2).
Adjusted R<sup>2</sup> for model B1: -0.049.
<sup>a</sup> defined as ferritin <13ng/ml. Reference group: ferritin ≥13ng/ml

## Discussion

In relation to our first hypothesis, vitamin D deficiency and insufficiency were strikingly prevalent in the migrant population when compared to the white British and sedentee women in Bangladesh, with 29% of the migrants (14 out of 49) being deficient (25(OH)D<25nmol/L). These women were therefore at potential risk of bone disorders, ranging from fractures and lower bone mineral density [15] to serious conditions such as osteomalacia [9].

To date, there is only one other study focusing on vitamin D in British-Bangladeshi adults [43], where mean 25(OH)D was found to be 44.0nmol/L, higher than our own findings of 32.2nmol/L. This could be due to the different immunoassay used [40], or may indicate that vitamin D status in British-Bangladeshis has become worse. A study in British-Bangladeshi children [44] found similar levels of 25(OH)D to the prior study in adults (mean 42nmol/L), as well as a strong association between low 25(OH)D and low iron status. We also found an association between low 25(OH)D and low iron status; however, we cannot determine the direction of causality from cross-sectional data, and it is possible that both are attributable to a third unknown factor. Our study has also replicated previous findings showing an association between lower vitamin D levels and higher BMI [27]. Aside from these earlier analyses of British-Bangladeshis, other studies exploring vitamin D deficiency in the wider British South Asian population have mostly shown similar trends to the study presented here [4,5].

There was a sharp contrast in vitamin D levels between the British-Bangladeshi migrants and the other groups: both the white British women and sedentee Bangladeshis were sufficient (on average), while fewer women in these comparison groups were deficient when compared to the Bangladeshi migrant group. These results replicate findings from previous studies [32,45], although our sedentee sample had higher 25(OH)D levels compared to other findings [6,46]. This is unlikely to be due to month of sampling; previous studies were performed in April-July, months which have a similar UV index to the period of our study (March and April) [47]. The higher levels may be due to the higher socioeconomic class of the participants in our study, with some of the effect attributable to different clothing and working practices, and the effect of these on exposure to sunlight and, therefore, vitamin D synthesis. It appears that in Bangladesh a high proportion of women are vitamin D insufficient, but a smaller proportion are deficient compared to the South Asian community in Britain.

We anticipated replicating the effects of previous studies showing significantly lower vitamin D levels in winter vs summer [4]. This was upheld for the white British women. Winter 25(OH)D was lower than summer 25(OH)D in the Bangladeshi migrant women, but not significantly so. This could be because dark-skinned migrants experience less variation in vitamin D status across seasons, as has been shown elsewhere [45].

Since the migrant and sedentee Bangladeshis came from the same social class, region and family background in Sylhet, Bangladesh, the main factors causing drastically lower levels of vitamin D in the migrant Bangladeshis are likely to be those associated with their northerly migration resulting in reduced UV exposure, particularly during the winter months [4], conforming to the notion of mismatch in evolutionary medicine. Lack of exposure to sunlight may be exacerbated by clothing worn by Bangladeshi women, which generally tends to cover them from neck to foot and may include a hijab. Cultural traditions may also affect how frequently women venture outside the home depending on their levels of acculturation and socioeconomic status. In addition, the population in question also transitioned from being middle-class in Sylhet to one of the poorest socioeconomic groups in the UK [48], and experienced many lifestyle changes, including dietary and working practices that may also play a role in vitamin D status [49].

### Limitations of the Study

Analyses were limited to available data from a study originally designed to examine reproductive ageing. For example, there was no specific question on vitamin D supplementation, or vitamin D dietary intake in the questionnaire. Moreover, the sample size of each group in this study was relatively small, which limited some of the statistical analyses.

Only one serum sample was taken from each participant. A longitudinal study would have been more robust, with multiple samples from the same participants over a period of time, preferably a year or more [4,45], to allow for comparison of seasonal variation in the same participants. The seasonal data were also constrained because the sedentee samples were taken in March and April due to the logistics of the original study, although this was less relevant given that UV radiation levels do not vary as much across the year in Bangladesh. Further research is needed to analyze seasonal variation in 25(OH)D among Bangladeshi sedentees.

## Conclusions and Implications

In the present study, a high proportion of Bangladeshi migrant women were mismatched to their current environment in that 29% were deficient in vitamin D, while a further 65.3% were insufficient in vitamin D, and only 6.1% were sufficient. If the study translates more widely to British-Bangladeshi migrant women aged 35-39, approximately 94% - many thousands of women - may be at increased risk of negative health outcomes, such as fractures, poor musculoskeletal health and perhaps cancer and cardiovascular disease especially in later life, as well as potentially higher morbidity and mortality from COVID-19. In contrast, >50% of white British and Bangladeshi sedentee women were sufficient in Vitamin D, with <6% of each group being deficient. Our study also replicated results showing seasonal variation in vitamin D levels in the white subjects, and lower but not significantly lower levels in winter in the Bangladeshi migrants [45].

As indicated in prior research, regular supplementation of vitamin D should be more widely recommended for British-Bangladeshis. This could be achieved through either increased dietary intake of vitamin D-rich foods or through direct supplementation, such as daily or weekly tablets of cholecalciferol [34]. Supplementation would be a relatively cost-effective method of improving health outcomes in this sub-group in the UK which already suffers from poorer health than the general population [50]. Such public health measures could yield tangible health benefits for thousands of British-Bangladeshis.

## Data Availability

Data held by University of Durham

## Acknowledgements

The authors’ contributions are as follows: LLS, SM, KB, LM, TS, RG, OC and GB were responsible for the data collection. NS, GB and LLS contributed to the study design and data analyses. NS wrote the manuscript, with GB and LLS providing guidance and review.

We thank all the women who participated in the study and particularly the Bengali Women’s Health Project for assistance. We are also thankful for the help of Susmita Paul Jamy, Sunali Biswas, Nusrath Jahan, Mahbuba Bari Shikder, Shamima Begum, Sayema Islam Shetu, Sufia Sultana, Rumana Begum, Kanij Fathema, Salma Begum, Khaza Nazim Uddin, and Tamanna Sultana.

---

**Table S1:**
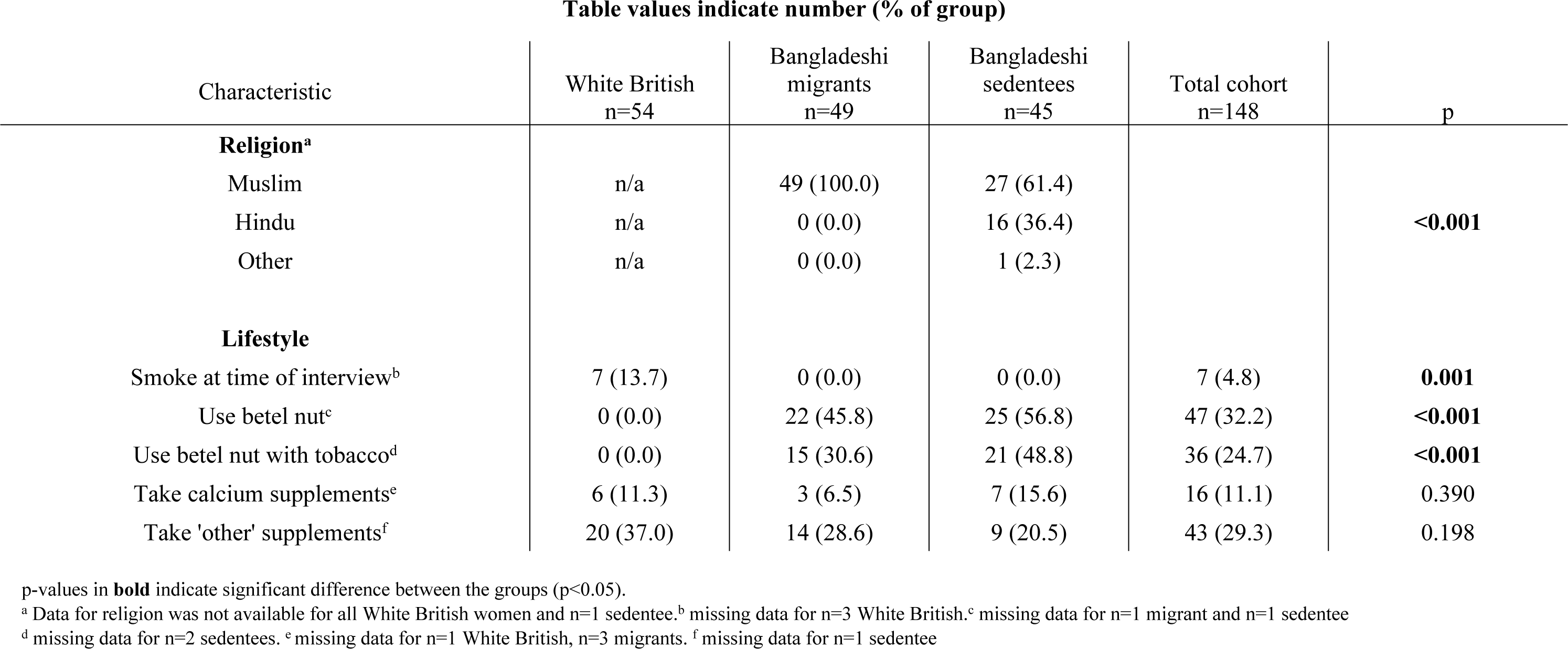
Other characteristics of the cohort.

**Table S2:** Season of serum sampling in the White British and Bangladeshi Migrant groups.

|  | 'Summer' (May-Oct) | 'Winter' (Nov-Apr) | Total |
| --- | --- | --- | --- |
| White British | 38 (70.4%) | 16 (29.6%) | 54 |
| Bangladeshi migrant | 33 (67.3%) | 16 (32.7%) | 49 |
| Total | 71 (68.9%) | 32 (31.1%) | 103 |

Fisher's exact test (2-sided) p=0.832

**Table S1:**
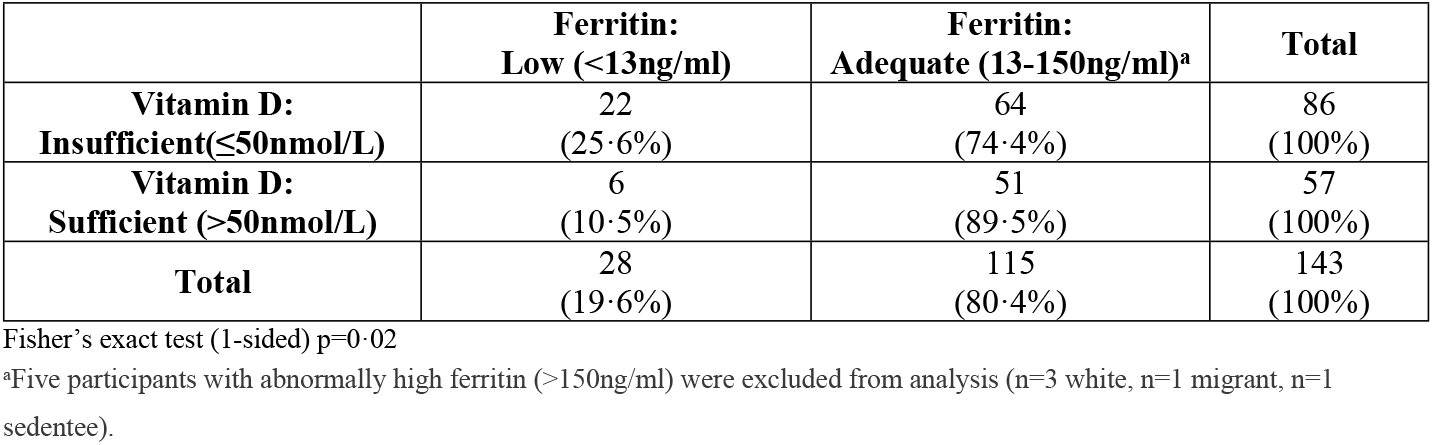
Vitamin D insufficiency and low iron status.

Women who were 25(OH)D insufficient (total n=89) had a significantly higher BMI than those who were ‘sufficient’ (n=59; mean BMI in insufficient group 26.6 vs 24.7 in sufficient group, p=0.009). When women were grouped by BMI into ‘healthy/underweight’ (<25) and ‘overweight/obese’ (≥25) the result was similar: mean 25(OH)D in the former category was 49.8nmol/L compared to 43.4nmol/L in the latter (p=0.043). Vitamin D levels were significantly negatively associated with increased tricep skin fold thickness and increased BMI but not with increased arm circumference. This remained true when these variables were analysed again, looking at mean values in women grouped into ‘insufficient’ (in vitamin D) vs ‘sufficient’ (Tables S3 and S4).

**Table S4:** Anthropometric measures and association with vitamin D as a continuous variable.

| Variable | Pearson correlation | p-value |
| --- | --- | --- |
| BMI | -0.172 | **0.037** |
| WHR<sup>a</sup> | -0.155 | 0.063 |
| Tricep skin fold thickness<sup>b</sup> (cm) | -0.239 | **0.004** |
| Arm circumference<sup>c</sup> (cm) | -0.116 | 0.169 |

Values in **bold** indicate significance ( $p=0.05$ )
<sup>a</sup> missing data for n=3 whites, n=1 migrant
<sup>b</sup> missing data for n=2 whites, n=1 migrant
<sup>c</sup> missing data for n=5 whites, n=1 migrant

**Table S5:**
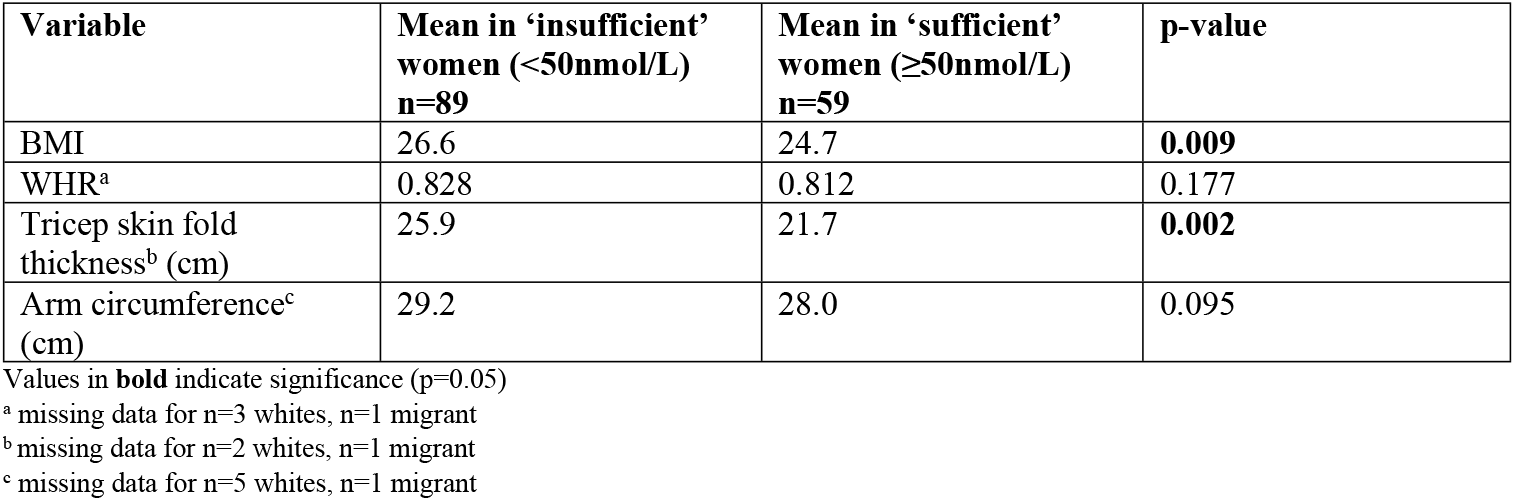
Anthropometric measures and association with insufficiency vs sufficiency status.

## Other cohort characteristics and vitamin D

There were no significant associations between age and Vitamin D when analysing the entire sample, or within each group separately. Educational status (defined by number of years of education, grouped into <10 years or ≥10 years) appeared to be significantly associated with concentrations of 25(OH)D (mean in <10 years education=42.8nmol/L, ≥10 years=50.5nmol/L, p=0.017). However, as there were many more migrants compared to whites in the <10 years education group, linear regression also adjusting for group was performed. This showed that there was no significant association between educational status and vitamin D (p=0.640), and that the previous association had 25 of 25 been due to education acting as a proxy for group status. On the adjusted education status scale, devised to account for the fewer years of education in the Bangladeshis, there was also no association with vitamin D (p=0.313).

There was no association between religion and mean vitamin D in the sedentees (this was not analysed in whites due to lack of data on religion, and all of the migrants were Muslim). The 17 non-Muslims had a similar mean 25(OH)D to the Muslims (51.3 vs 50.4nmol/L respectively, p=0.829) and were no more likely to be insufficient in vitamin D (p=0.535).

In addition, no linear association was found between length of time since migration and vitamin D 43 (p=0.638).

Use of either cigarettes or betel nut with tobacco was not significantly associated with Vitamin D levels (p=0.426). Use of either calcium or vitamin supplements was also not associated with significantly higher Vitamin D (p=0.786). These results are shown in table S5.

**Table S6:** Associations between lifestyle factors, migration status and vitamin D.

|  |  | Mean<br>25(OH)D<br>(nmol/L) | p-value |
| --- | --- | --- | --- |
| Smoking or use of tobacco with betel nut<sup>a</sup> | Either<br>n=43 | 48.5 | 0.426 |
|  | None<br>n=98 | 45.7 |  |
| Supplements: combined<sup>b</sup> | Take any<br>n=46 | 46.6 | 0.786 |
|  | Take none<br>n=97 | 45.7 |  |
| Supplements: calcium<sup>c</sup> | Yes<br>n=16 | 48.4 | 0.632 |
|  | No<br>n=128 | 46.0 |  |
| Supplements: other vitamins and minerals<sup>d</sup> | Yes<br>n=43 | 46.7 | 0.769 |
|  | No<br>n=104 | 45.7 |  |

<sup>a</sup> missing data for n=3 white British, n=2 migrants and n=2 sedentees
<sup>b</sup> missing data for n=1 white British, n=3 migrants, n=1 sedentee
<sup>c</sup> missing data for n=1 white British, n=3 migrants
<sup>d</sup> missing data for n=1 sedentee

